# SARS-CoV-2 seroprevalence survey estimates are affected by anti-nucleocapsid antibody decline

**DOI:** 10.1101/2020.09.28.20200915

**Authors:** Shelly Bolotin, Vanessa Tran, Selma Osman, Kevin A. Brown, Sarah A. Buchan, Eugene Joh, Shelley L. Deeks, Vanessa G. Allen

## Abstract

We analyzed 21,676 residual specimens from Ontario, Canada collected between March-August, 2020 to investigate the effect of antibody decline on SARS-CoV-2 seroprevalence estimates. Testing specimens orthogonally using the Abbott (anti-nucleocapsid) and then the Ortho (anti-spike) assays, seroprevalence estimates ranged from 0.4%-1.4%, despite ongoing disease activity. The geometric mean concentration (GMC) of antibody-positive specimens decreased over time (p=0.015), and the GMC of antibody-negative specimens increased over time (p=0.0018). The association between the two tests decreased each month (p<0.001), suggesting anti-N antibody decline. Lowering the Abbott index cut-off from 1.4 to 0.7 resulted in a 16% increase in positive specimens.

## Background

Severe acute respiratory syndrome coronavirus 2 (SARS-CoV-2), which causes coronavirus disease 2019 (COVID-19), emerged as a novel pathogen in late 2019. By mid-September, 2020 it has resulted in over 28 million cases and nearly one million deaths [1]. In Ontario, Canada, the first pandemic wave peaked in mid-April [2]. This was followed by lower but sustained disease activity between June and August, with elevated case counts in various hot-spots in the province. By September 22, 2020 there were approximately 48,000 cases reported provincially for a population of 14.8 million residents [2].

As part of Ontario’s pandemic response, Public Health Ontario (PHO) initiated a SARS-CoV-2 serosurveillance program to estimate the proportion of the population that has been infected, identify demographic risk factors for infection, and enhance the pandemic response [3]. The program utilizes residual specimens submitted to the PHO Laboratory from March, 2020 onwards. We tested specimens using an orthogonal approach in order to minimize false positives, particularly when population prevalence is low, in line with a recommendation by the Centers for Disease Control and Prevention [4]. Despite ongoing disease activity [2], we were surprised to discover that there were no substantial changes in Ontario seroprevalence estimates over the first five serosurveys conducted from March to August, 2020, with very low seroprevalence estimates throughout this period [3 and unpublished]. Recent studies by others have reported declining antibody levels in confirmed SARS-CoV-2 cases. However, these findings have generally been based on symptomatic, confirmed individuals or small groups of asymptomatic cases, with limited reporting of antibody decline at the population level [5–7]. We therefore investigated population-level antibody decline using Ontario serosurveillance data.

## Methods

### Specimen source

We conducted five cross-sectional SARS-CoV-2 serosurveys (March 27 – April 30; May 24 – 31; June 5 – 30; July 4 – 31; August 1-21, 2020), using residual sera, plasma and blood specimens remaining after routine clinical testing at the PHO Laboratory. Specimens were selected to represent both sexes, all age groups and all geographic regions in Ontario.

### Laboratory testing

Using an orthogonal testing approach, we first tested specimens with the Abbott Architect SARS-CoV-2 IgG test (Abbott Laboratories, USA), which detects anti-nucleocapsid (N) antibodies, then tested positive samples with the Ortho-Clinical Diagnostics VITROS Anti-SARS-CoV-2 IgG test (Ortho-Clinical Diagnostics, Inc., USA), which detects anti-spike (S) antibodies [8]. We used the manufacturers’ recommended index value cut-off of 1.4 and 1.0 for the Abbott and Ortho assays, respectively. Specimens that were positive using both tests were considered positive for SARS-CoV-2 antibodies. To explore the effect of lowering the Abbott index value on population seroprevalence, we analyzed a subset of specimens with an index value of 0.7, otherwise maintaining the same orthogonal approach as above.

### Statistical analysis

To investigate changes in the average antibody concentration over time, we calculated the geometric mean concentration (GMC) of serology specimens for each collection period. Since GMC cannot be calculated with zero values, we first replaced zero index values with a value of 0.01/ √2 [9], with 0.01 being the lowest non-zero value reported by the Abbott instrument. We used linear regression to assess the trend in GMC across months and truncated linear regression to examine the association between the Abbott and Ortho index values through time [10]. Truncation was used to account for potential bias introduced by using orthogonal testing since this analysis only includes samples where the Abbott result was ≥1.4. We fit one model for the overall association as well as one model for each of the five monthly serosurveys. To investigate the relationship between the Abbott and Ortho results over time, we fit a regression model with the statistical interaction between date and the slope to measure the change in slope through time.

### Ethics

Ethics approval for the serosurveys was granted by the PHO Ethics Review Board

## Results

We tested a total of 21,676 specimens over the five time periods. Of these, 3/827 (0.4%, 95% confidence interval (CI) 0.1, 1.1) were positive in March-April, 15/1,061 (1.4%, 95% CI 0.7, 2.1) in May, 79/7,023 (1.1%, 95% CI 0.9, 1.4) in June, 70/7,001 (1.0%, 95% CI 0.8, 1.2) in July, and 64/5,764 (1.1%, 95% CI 0.8, 1.4) in August (Table 1). Overall, 318 specimens were positive for SARS-CoV-2 antibodies using the Abbott assay, of which 231 (72.6%) were also positive using the Ortho assay.

**Table 1:** Serology test results and geometric mean concentration of Ontario serosurveillance specimens, March – August, 2020.

| Test results | Specimen collection period |  |  |  |  |
| --- | --- | --- | --- | --- | --- |
|  | March - April | May | June | July | August |
| **Abbott** |  |  |  |  |  |
| Positive n/N | 5/827 | 20/1,061 | 106/7,023 | 97/7,001 | 90/5,764 |
| Positive AI GMC (95% CI)* | 3.4 (1.9, 6.0) | 3.7 (3.0, 4.5) | 3.7 (3.4, 4.1) | 3.3 (3.0, 3.7) | 3.1 (2.8, 3.4) |
| Negative n/N | 822/827 | 1,041/1,061 | 6,917/7,023 | 6,904/7,001 | 5,674/5,764 |
| Negative AI GMC (95% CI)* | 0.034 (0.032, 0.037) | 0.032 (0.031, 0.034) | 0.033 (0.032, 0.033) | 0.034 (0.033, 0.035) | 0.035 (0.034, 0.035) |
| **Ortho<sup>a</sup>** |  |  |  |  |  |
| Positive n/N | 3/5 | 15/20 | 79/106 | 70/97 | 64/90 |
| Positive AI GMC (95% CI) | 22 (13, 38) | 15 (11, 22) | 18 (15, 21) | 16 (13, 19) | 15 (13, 18) |
| Negative n/N | 2/5 | 5/20 | 27/106 | 27/97 | 26/90 |
| Negative AI GMC (95% CI) | 0.046 (0.0055, 0.38) | 0.047 (0.012, 0.18) | 0.029 (0.020, 0.042) | 0.021 (0.015, 0.030) | 0.028 (0.019, 0.042) |
| **Orthogonally positive** |  |  |  |  |  |
| n/N | 3/827 | 15/1,061 | 79/7,023 | 70/7,001 | 64/5,764 |
| % (95% CI) | 0.4 (0.1, 1.1) | 1.4 (0.7, 2.1) | 1.1 (0.9, 1.4) | 1.0 (0.8, 1.2) | 1.1 (0.8, 1.4) |

<sup>a</sup> = only Abbott-positive specimens were tested; GMC = geometric mean concentration; AI = antibody index; CI= confidence interval; \* = statistically significant trend across sampling periods

To explore whether SARS-CoV-2 antibody decline could account for the lack of increase in seroprevalence estimates over time, we first compared the GMCs for each sampling period. GMCs of specimens that were positive by the Abbott assay (n=318), which was the only assay with which all specimens were tested, were relatively stable for our first three sampling periods. However, a decline was observed from 3.7 (95% CI 3.4, 4.1) in June to 3.3 (95% CI 3.0, 3.7) in July, and then to 3.1 (95% CI 2.8, 3.4) in August (linear regression p for all sampling periods =0.015) (Table 1). Conversely, the GMC of specimens that tested negative by the Abbott assay (n=21,358) increased incrementally from May to August (linear regression p for all sampling periods =0.0018). The statistically significant decrease and increase in GMCs over time for both Abbott positive and negative specimens, respectively, suggests that population-level SARS-CoV-2 antibodies has declined over time. The GMC of positive and negative specimens using the Ortho assay fluctuated over time (Table 1), but changes were not statistically significant (linear regression p=0.29 and 0.35, respectively).

To explore how antibody decline might impact our seroprevalence estimates, we examined the relationship between the Abbott and Ortho assay results over time for the 318 specimens that were tested using both assays. Overall, there was a strong association between the assays, such that each Ortho index value increase of 10 was associated with an Abbott index value increase of 1.4 (95% CI 1.2, 1.7). Additionally, we found that the magnitude of the association decreased over time (Figure 1). Specifically, we found that in April, each Ortho index value increase of 10 was associated with an Abbott index value increase of 2.5 (95% CI 0.005, 5.1), while in June the association was 1.5 (95% CI 1.2, 1.8), and in August it was 1.3 (95% CI 0.4, 2.3). Overall, there was an index value decrease of 0.3 (95% CI 0.1, 0.5) in the slope of the association for each additional month since April (p<0.001).

**Figure 1.**
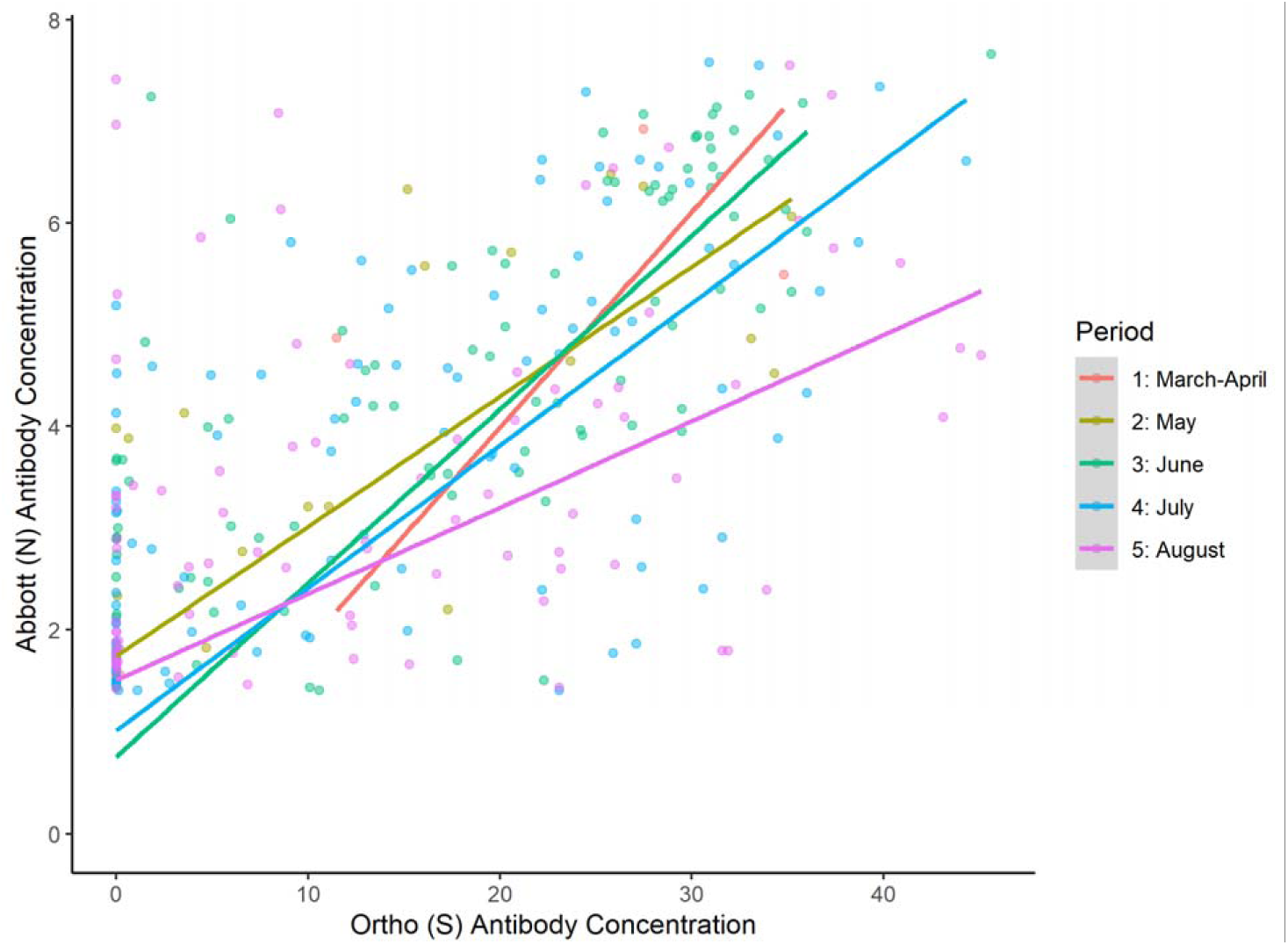
The association between Ortho and Abbott antibody index values by month, March to August, 2020.

Our observed trend of anti-N antibody decline using the Abbott assay has potential implications on seroprevalence estimates, because using our orthogonal approach specimens that test negative using the Abbott assay would not be subsequently tested using the Ortho assay, and would be deemed negative. To further explore this, we tested a subset of specimens collected in August, 2020 (N = 4,166) with the same orthogonal approach, but lowered the Abbott index value to 0.7 to account for anti-N antibody decline. While using a cut-off of 1.4 resulted in 43/4,166 specimens (1.0%) that were positive by both assays, lowering the index resulted in seven additional positive specimens, for a total of 50/4,166 (1.2%).

## Discussion

Our investigation of population-level antibodies to SARS-CoV-2 revealed a decline in population level anti-N antibodies detected by the Abbott Architect SARS-CoV-2 IgG assay, over a five month period. This finding has implications for serosurveys that use the Abbott Architect assay or other anti-N assays, alone or as part of an orthogonal approach. Our results suggest that the anti-N antibody decline is substantial enough to affect the results of population seroprevalence surveys, especially in high prevalence settings. Due to overall low seroprevalence in Ontario, our observed antibody decline represented only a small change in estimates, from 1.0% to 1.2%. However, lowering the Abbott index cut-off resulted in a 16% increase in positive specimens, which would make a substantial difference in jurisdictions where population seroprevalence was higher. Since seroprevalence data are often used to estimate the proportion of individuals infected, including those who are not captured as cases through RT-PCR testing, antibody decline may also result in an underestimate of SARS-CoV-2 infections at the population level.

Others investigating SARS-CoV-2 antibody decline have reported conflicting results. Assessing individual immune responses, Long et al. [6] found decline in both anti-N and anti-S IgG antibody levels by eight weeks after hospital discharge. Conversely, Wu and colleagues reported stable anti-S and anti-N IgG activity six months post-infection [11]. Similarly, an analysis of >1,000 specimens from Icelandic patients using various assays directed against both anti-N and anti-S antibodies found durable IgG responses four months after diagnosis [5]. Exploration of antibody decline at the population level is less common. However, Public Health England recently reported an increase in equivocal serosurveillance results using an anti-S IgG assay [12]. It is likely that contrasting study results are related to differences between the populations sampled, time of sampling in relation to onset of infection, as well as the variety of assays used [5].

There are several modifications that can be made to SARS-CoV-2 serosurveillance testing protocols to address the issue of antibody decline, while still maintaining an orthogonal approach. We have shown here that lowering the antibody index value of the Abbott anti-N assay to increase sensitivity to account for potential antibody decline would result in a higher proportion of positive specimens. However, we recognize that this would change the test characteristics markedly. Using sera from SARS-CoV-2 RT-PCR positive patients, a study by Bryan and colleagues determined that lowering the Abbott assay index cut-off from 1.4 to 0.7 resulted in test sensitivity (≥7 days from symptom onset) rising from 84.2% to 88.0%, and test specificity decreasing from 100.0% to 99.6% [13]. However, retaining an orthogonal approach may mitigate the reduced specificity. Alternatively, switching to a different anti-N assay may alleviate the issue of antibody decline. Although anti-N antibodies have been shown to decline faster than anti-S antibodies, the magnitude of decline seems to be somewhat assay-specific, and therefore switching anti-N assays may result in more durable serology measures [14]. Consideration should be given to the portion of the N antigen that is used in serology assays, as this may impact the ability to detect a durable antibody response.

Our analysis included >21,000 specimens over five months, representing SARS-CoV-2 unexposed individuals as well as infected individuals of all ages, who likely experienced a spectrum of SARS-CoV-2 disease symptoms and severity. This is a strength of our study, since many analyses of SARS-CoV-2 antibody decline to date have focused on individual humoral responses using specimens from symptomatic individuals who presented to medical care, or small groups of asymptomatic individuals [5–7]. While these studies are essential for characterizing individual immune responses to SARS-CoV-2, they have a limited ability to shed light on population-level antibody decline, which may have different kinetics than those observed in small groups of previously infected individuals [5]. However, our study was limited by the cross-sectional nature of our serosurveys. Since our specimens were de-identified, we could not follow individual antibody concentrations longitudinally to explore the determinants of antibody decline, such as disease severity, age or sex. This also limited our ability to explore the reasons behind the trend of increasing GMC in Abbott-negative specimens. While we suspect that the negative specimens included samples from infected individuals that declined into the non-reactive range of the test, our cross-sectional approach does not allow us to investigate this further. Lastly, due to our testing algorithm, only a subset of our specimens were tested using the Ortho assay. Changes in the Ortho GMC were not statistically significant, and we suspect this may be due to selection bias towards specimens with higher antibody concentrations in this group. Although we took this selection bias into account by using a truncated regression model to compare our two assays, it still limited our ability to characterize anti-S antibody levels over time.

As the COVID-19 pandemic progresses, it is becoming clear that although serology tests may not always correlate to virus neutralization [15], they are integral to understanding population exposure, since not every infected individual is tested and reported as a case. New solutions to mitigate the issue of antibody decline are a priority area of research.

## Data Availability

The datasets generated during and/or analysed during the current study are available from the corresponding author on reasonable request.

## Author contributions

All authors have contributed to this work and have approved the final version of the manuscript. SB conceived the idea for the study. SB, KB, SO, EJ and VT analyzed the data, and all authors provided subject-matter expertise with interpreting the data.

## References

1. Organization WH. Coronavirus disease (COVID-2019) situation reports [Internet]. 2020. Available from: https://www.who.int/emergencies/diseases/novel-coronavirus-2019/situation-reports

2. Ontario Agency for Health Protection and Promotion (Public Health Ontario). Ontario COVID-19 Data Tool. 2020;. Available from: https://www.publichealthontario.ca/en/data-and-analysis/infectious-disease/covid-19-data-surveillance/covid-19-data-tool

3. Ontario Agency for Health Protection and Promotion (Public Health Ontario). COVID-19 Seroprevalence in Ontario: March 27, 2020 to June 30, 2020 [Internet]. 2020. Available from: https://www.publichealthontario.ca/-/media/documents/ncov/epi/2020/07/covid-19-epi-seroprevalence-in-ontario.pdf?la=en

4. Centers for Disease Control and Prevention (CDC). Interim Guidelines for COVID-19 Antibody Testing [Internet]. 2020. Available from: https://www.cdc.gov/coronavirus/2019-ncov/lab/resources/antibody-tests-guidelines.html

5. Gudbjartsson DF, Norddahl GL, Melsted P, et al. Humoral Immune Response to SARS-CoV-2 in Iceland. N Engl J Med [Internet]. Massachusetts Medical Society; 2020;. Available from: https://doi.org/10.1056/NEJMoa2026116

6. Long Q-X, Tang X-J, Shi Q-L, et al. Clinical and immunological assessment of asymptomatic SARS-CoV-2 infections [Internet]. Nat. Med. 2020. Available from: https://doi.org/10.1038/s41591-020-0965-6

7. Patel MM, Thornburg NJ, Stubblefield WB, et al. Change in Antibodies to SARS-CoV-2 Over 60 Days Among Health Care Personnel in Nashville, Tennessee. JAMA [Internet]. 2020;. Available from: https://doi.org/10.1001/jama.2020.18796

8. U.S Food & Drug Administration. EUA Authorized Serology Test Performance [Internet]. 2020. Available from: https://www.fda.gov/medical-devices/emergency-situations-medical-devices/eua-authorized-serology-test-performance

9. Canales RA, Wilson AM, Pearce-Walker JI, Verhougstraete MP, Reynolds KA. Methods for Handling Left-Censored Data in Quantitative Microbial Risk Assessment. Schaffner DW, editor. Appl Environ Microbiol [Internet]. 2018; 84(20):e01203–18. Available from: http://aem.asm.org/content/84/20/e01203-18.abstract

10. Messner JW, Mayr GJ, Zeileis A. Heteroscedastic Censored and Truncated Regression with crch. R J. 2016; 8(1):173.

11. Wu J, Liang B, Chen C, et al. SARS-CoV-2 infection induces sustained humoral immune responses in convalescent patients following symptomatic COVID-19. medRxiv [Internet]. 2020; :2020.07.21.20159178. Available from: http://medrxiv.org/content/early/2020/07/24/2020.07.21.20159178.abstract

12. Public Health England. Weekly Coronavirus Disease 2019 (COVID-19) Surveillance Report [Internet]. 2020. Available from: https://assets.publishing.service.gov.uk/government/uploads/system/uploads/attachment_data/file/907954/Weekly_COVID19_Surveillance_Report_week_32_2.pdf

13. Bryan A, Pepper G, Wener MH, et al. Performance Characteristics of the Abbott Architect SARS-CoV-2 IgG Assay and Seroprevalence in Boise, Idaho. J Clin Microbiol [Internet]. 2020/05/10. Department of Laboratory Medicine, University of Washington School of Medicine, Seattle, WA. Department of Medicine, University of Washington School of Medicine, Seattle, WA. Vaccine and Infectious Disease Division, Fred Hutchinson Cancer Research Center,; 2020;. Available from: https://dx.doi.org/10.1128/jcm.00941-20

14. Muecksch F, Wise H, Batchelor B, et al. Longitudinal analysis of clinical serology assay performance and neutralising antibody levels in COVID19 convalescents. medRxiv [Internet]. 2020; :2020.08.05.20169128. Available from: http://medrxiv.org/content/early/2020/08/06/2020.08.05.20169128.abstract

15. Rijkers G, Murk J-L, Wintermans B, et al. Differences in Antibody Kinetics and Functionality Between Severe and Mild Severe Acute Respiratory Syndrome Coronavirus 2 Infections. J Infect Dis [Internet]. 2020; 222(8):1265–1269. Available from: https://doi.org/10.1093/infdis/jiaa463

